# Steroid Metabolome Profiling Identifies a Unique Androgen Hormone Signature Associated with Endometriosis

**DOI:** 10.1101/2025.05.24.25328281

**Authors:** Ioannis Simitsidellis, Rebecca Ainslie, Angela Taylor, Lorna C. Gilligan, Fozia Shaheen, Samira Blanke, Craig Anderson, Dharani Hapangama, Wiebke Arlt, Andrew W Horne, Philippa T.K. Saunders, Douglas A. Gibson

**Affiliations:** Centre for Reproductive Health, Institute for Regeneration and Repair, University of Edinburgh, Edinburgh BioQuarter, 4-5 Little France Crescent, Edinburgh EH16 4UU, UK; Metabolism and Systems Science, School of Medicine and Health, University of Birmingham, Birmingham, B15 2TT, UK; School of Mathematics and Statistics, University of Glasgow, 132 University Pl, Glasgow G12 8TA, UK; Department of Women’s & Children’s Health, Institute of Life Course and Medical Sciences, University of Liverpool, UK; Institute of Clinical Sciences, Imperial College London, London W12 0HS, UK; MRC Laboratory of Medical Sciences, London, W12 0HS, UK

## Abstract

Endometriosis is a chronic, hormone-dependent condition that affects 190 million women worldwide. There are no validated biomarkers for endometriosis and this delays diagnosis and treatment.

We performed serum steroid metabolome profiling in healthy controls (n=57) and women with laparoscopically-confirmed endometriosis (n=159) using liquid chromatography-tandem mass spectrometry. Women with endometriosis had a distinct steroid signature characterised by increased concentrations of classic and 11-oxygenated androgens, and altered metabolism associated with 11-ketotestosterone production.

Metabolomic data were used to generate a supervised machine learning model to predict diagnostic outcome. ROC curve analysis demonstrated robust discrimination between healthy controls and endometriosis patients (AUC=0.99) with 96.84% positive-, and 92.86% negative-predictive power. Data were partitioned into train and validation groups, and a refined model identified >95% of endometriosis patients in a blinded sample set.

These data reframe endometriosis as an androgen-dominant condition and present a unique opportunity to develop novel diagnostic approaches using 11-oxygenated androgens as biomarkers.

## Introduction

Endometriosis is a chronic, hormone-dependent condition that affects an estimated 190 million women worldwide ^1^. It is characterised by the presence of ‘endometrial-like’ tissue (referred to as ‘lesions’) outside the uterus, most commonly within the pelvic peritoneal cavity. Women experience life-altering symptoms including chronic pelvic pain and infertility. In addition to impacts on quality of life, endometriosis also has serious financial consequences limiting health, wellbeing and life potential ^2^.

Diagnosis is a key clinical challenge in endometriosis, which prolongs suffering and delays treatment. Internationally, overall time to diagnosis ranges from 5 to 12 years^3^, with a median delay of 8 years reported in the UK^4^. Establishing a correct diagnosis of endometriosis is often problematic because the presenting symptoms are associated with other conditions and this makes clinical recognition of endometriosis challenging ^5,6^. Thus, the diagnosis of endometriosis usually involves a combination of clinical evaluation and imaging techniques ^7^. However, the gold standard for diagnosing endometriosis, as recommended by international guidelines, is laparoscopic visualisation of the lesions performed by specialised gynaecologists ^8–10^. Endometriosis is broadly categorised into three subtypes: superficial peritoneal, ovarian and deep. Nevertheless, it is most commonly described according to the revised scoring system of the American Society for Reproductive Medicine (rASRM) which ranges from Stage I to Stage IV based on the location and extent of lesions, as well as adhesions, visualized at surgery ^11^. Although widely adopted, this surgical classification does not correlate with symptom severity ^12^. Additionally, diagnostic laparoscopy is invasive, costly, and carries significant risks, including injury to pelvic organs ^13^. Non-invasive diagnostic approaches for endometriosis could provide earlier definitive diagnosis and eliminate surgery-associated complications. However, unlike other similarly common diseases, there are no known biomarkers for endometriosis.

Research focused on identifying endometriosis biomarkers has been hampered by low-powered studies, inter-study variability and by the identification of markers with low predictive value ^14^. Until now, there has been no single blood or urinary biomarker approach that has been validated ^15^. Recent attention has focussed on screening salivary miRNAs ^16^ and the serum proteome as potential diagnostic approaches but independent validation studies are yet to be reported ^17,18^. A common limitation of endometriosis biomarker studies is that they seek to screen out differential targets without consideration of the underlying biology, limiting their utility for further development as therapeutic targets and prognostic markers.

Endometriosis is a hormone-dependent disorder but utilising endocrine differences for diagnostic purposes is challenging. Estrogens are considered the main disease driver due to their established roles in promoting inflammation, angiogenesis and proliferation of endometriosis lesions ^19^. Although estrogen concentrations are increased within lesions there are no reported differences in circulating estrogens in women with endometriosis ^20,21^. Furthermore, estrogen concentrations are highly variable across the menstrual cycle, limiting their potential utility as diagnostic biomarkers. In contrast, androgens, which can be derived from both the ovary and the adrenal, have limited cyclical variation in women. Classic androgens, such as testosterone (T) and dihydrotestosterone (DHT), regulate various cellular processes implicated in endometriosis, including proliferation, tissue remodeling, and inflammation ^22^. Testosterone concentrations are increased in endometriosis lesions and this is associated with changes in androgen-regulated genes ^23^.

Recent studies suggest that adrenal-derived 11-oxygenated androgens are important components of the androgen pool, particularly in women ^24^, but little is known about the potential contribution of 11-oxygenated androgens to endometriosis pathophysiology. 11KT circulates at concentrations equivalent to or greater than T in women and has similar androgenic activity, consistent with a potential role in regulating androgen-dependent processes ^25^. 11-oxygenated androgens are now recognised for their roles in endocrine disorders such as polycystic ovarian syndrome (PCOS) ^26^ but whether they also play a role in endometriosis is yet to be determined.

Given the potential importance of androgens to endometriosis pathophysiology and the limited data on profiling of androgen metabolites, we sought to define the androgen metabolome in endometriosis and investigate its potential utility as a diagnostic approach. Serum samples from women with laparoscopically-confirmed endometriosis (n=159) and healthy controls (n=61) were profiled using a liquid chromatography–tandem mass spectrometry (LC-MS/MS) assay to simultaneously measure classic and 11-oxygenated androgens. Women with endometriosis had a distinct steroid signature characterised by increased concentrations of classic and 11-oxygenated androgens and altered metabolism associated with 11KT production. In predictive statistical models, 11KT and related metabolites demonstrated significant potential as diagnostic biomarkers and could robustly distinguish between healthy controls and women with endometriosis. These data reframe endometriosis as an androgen-dependent disorder and highlight the potential for 11-oxygenated androgens to be used as diagnostic biomarkers.

## Results

### Endometriosis is associated with an altered androgen hormone signature

Androgens and their precursors were quantified using LC-MS/MS in serum samples from healthy controls (HC; n=57) and women with endometriosis (ENDO; n=159). The concentrations of ‘classic’ androgens; DHEA, A4, T and DHT and 11-oxygenated androgens; 11OHA4, 11OHT, 11KA4 and 11KT were determined (Figure 1 and Table 1).

**Figure 1.**
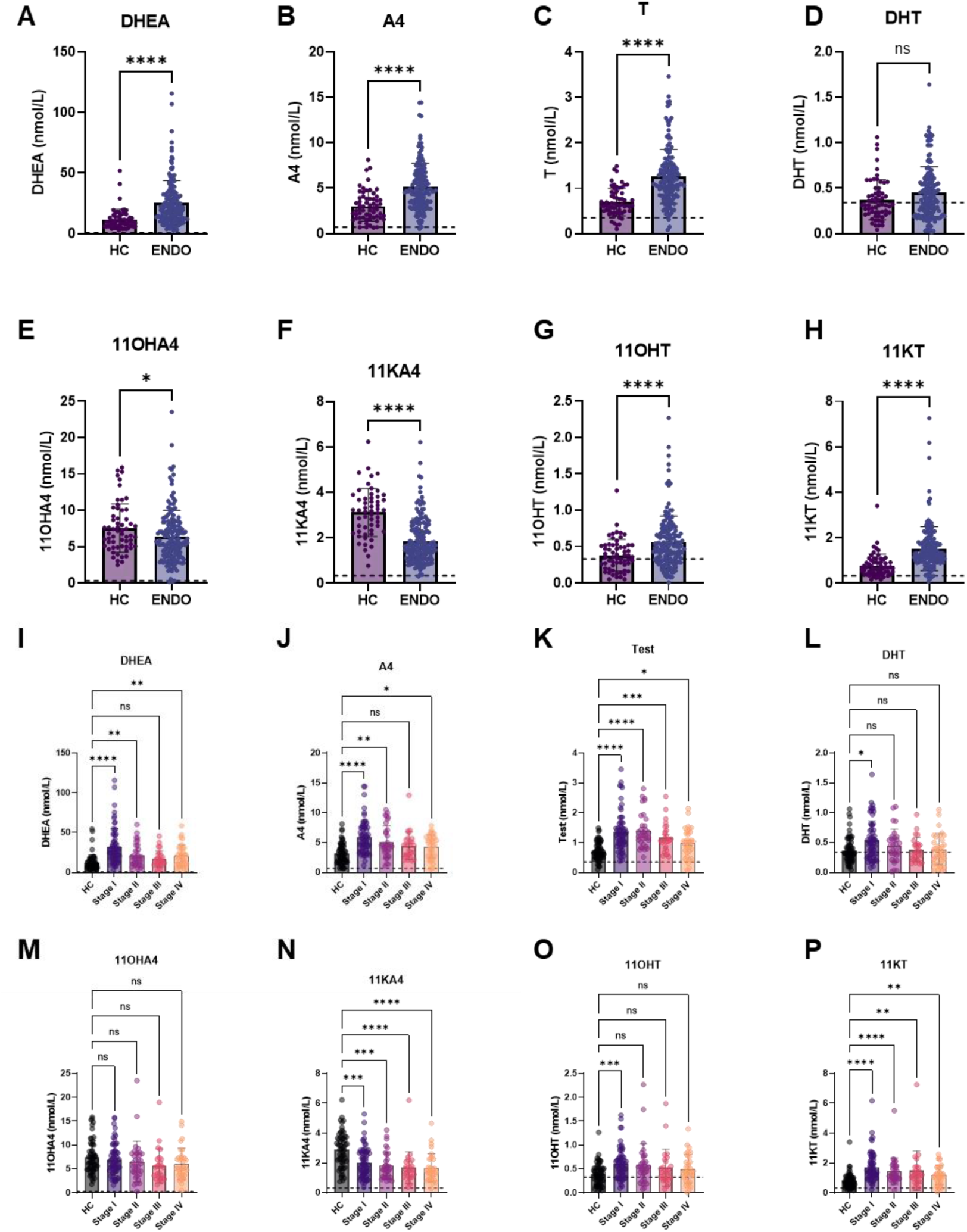
Serum concentrations of classic androgens and 11-oxygenated androgens in healthy controls (HC) or women with endometriosis (ENDO). Serum concentrations of DHEA, A4, T, and 11OHA4, 11KA4, 11OHT, and 11KT were by LC-MS/MS. DHEA, A4, T, were significantly elevated in ENDO compared to HC (**A**-**C**). Concentrations of DHT did not differ between groups (**D**). 11-oxygenated androgen precursors, 11OHA4 (**E**) and 11KA4 (**F**), were decreased in ENDO, while potent 11-oxygenated androgens, 11OHT (**G**) and 11KT (**H**) were significantly elevated in ENDO. ENDO patients were stratified according to rASRM stage (**I**-**P**). DHEA and A4 were significantly elevated in Stage I, II and IV endometriosis (**I**, **J**). T was elevated in all endometriosis stages (**K**), and DHT was significantly elevated in Stage I endometriosis (**L**). Concentrations of 11OHA4 did not significantly differ between groups (**M**) while 11KA4 was decreased in all stages (**N**). In contrast, 11OHT was significantly elevated in Stage I endometriosis (**O**) while 11KT was elevated in all endometriosis stages (**P**). Median serum steroid concentrations (25th-75th centile range; nmol L–1) for each stage (I). Error bars represent the standard deviation of the mean. Dotted lines denote LLOQ for each analyte. Undetected steroids or those detected below LLOQ were replaced by 0.5×LLOQ for statistical purposes. Statistical comparison Mann Whitney U test or Kruskal-Wallis test with multiple comparisons. *p<0.05, **p<0.01, ***p<0.001 ****p<0.0001, ns – non significant.

**Table 1.**
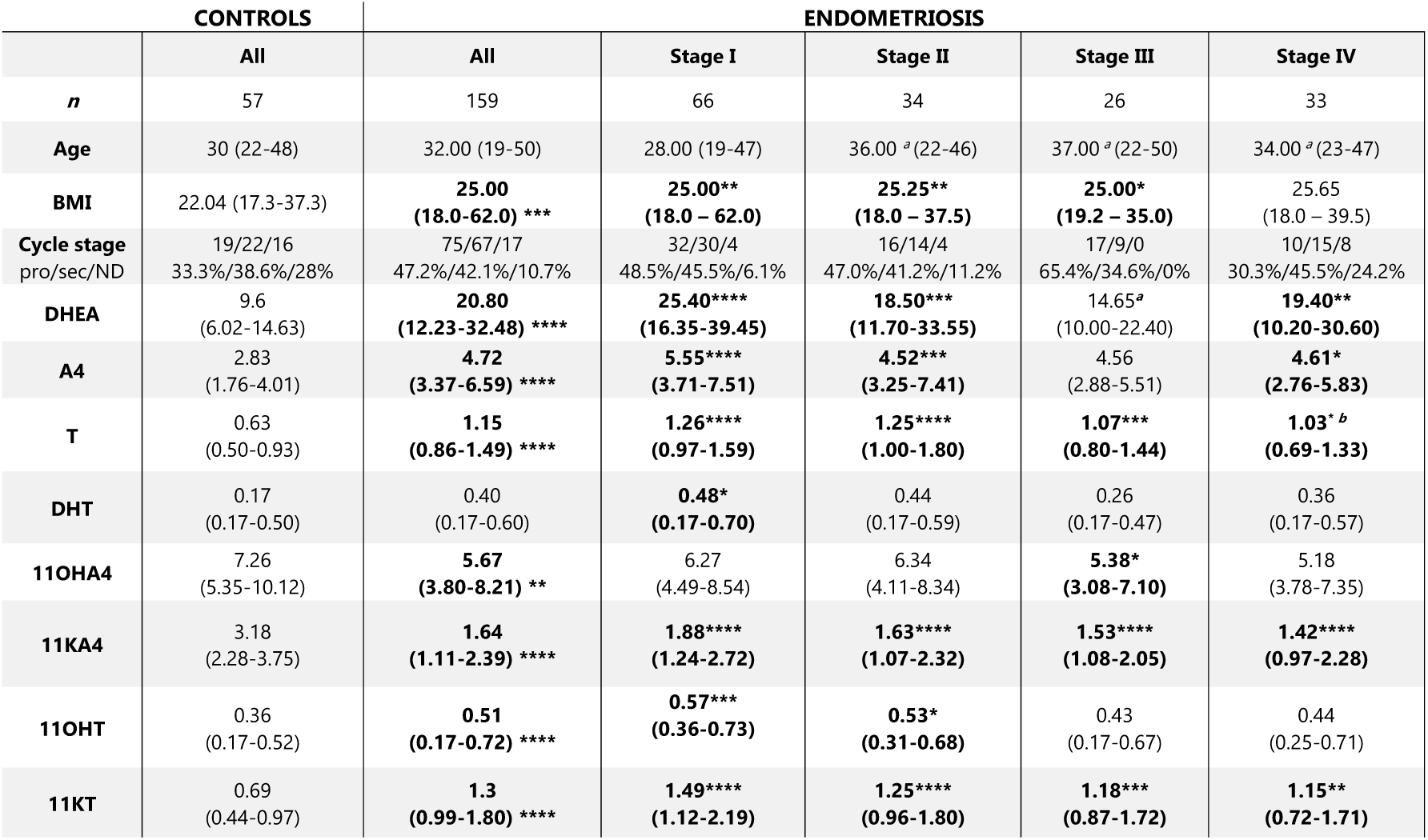
Serum steroid concentrations of classic and 11-oxygenated androgens in healthy controls and women with endometriosis. Age (years) median (min-max), BMI (kg/m2) median (min-max), serum steroid concentrations (median and 25th-75th centile range; nmol L–1). Values below the lower limit of quantification are indicated as < lower limit of quantification. Steroid concentrations below the LLOQ were replaced by 0.5×LLOQ for statistical purposes. P value vs healthy control; * p<0.05, ** p<0.01, *** p<0.001, **** p<0.0001. ^a^ vs Stage I, ^b^ vs Stage II.

DHEA (Figure 1A; p<0.0001), A4 (Figure 1B; p<0.0001) and T (Figure 1C; p<0.0001) were significantly higher in serum samples from women with endometriosis compared to healthy controls (Table 1 ‘All’). Concentrations of DHT were not significantly different between groups (Figure 1D). The 11-oxygenated androgen precursor metabolites, 11OHA4 (Figure 1E; p<0.05) and 11KA4 (Figure 1F; P<0.0001), were decreased, and the active 11-oxygenated androgens, 11OHT (Figure 1G; p<0.0001) and 11KT (Figure 1H; p<0.0001), were increased in women with endometriosis (Table 1 ‘All’). Consistent with previous reports ^27^, 11-oxygenated androgens did not differ by menstrual cycle phase (Supplementary Table 1). Neither classic nor 11-oxygenated androgen concentrations correlated with age, although 11OHT trended to positive correlation (r=0.1267, p=0.0636). Only 11KA4 negatively correlated with BMI (r=-0.2029, p=0.0037; Supplementary Table 2).

### Classic and 11-oxygenated androgens do not differ by rASRM stage

To assess whether androgen concentrations were associated with disease severity in endometriosis, data were stratified according to rASRM stage. Broadly, there were no significant differences in steroid concentrations between rASRM stages for any of the analytes assessed. Analytes that were increased in the unstratified group (ENDO/‘All’) were also increased in each rASRM stage compared to healthy controls (HC) (Figure 1 I-P; Table 1). Notably, median concentrations of DHEA (p<0.0001), A4 (p<0.0001) and T (p<0.0001) were highest in Stage I endometriosis and significantly higher than healthy controls (Figure 1 I-P; Table 1). Similarly, concentrations of 11OHT (p<0.001) and 11KT (p<0.0001) were highest in Stage I endometriosis and significantly higher than healthy controls (Figure 1I-P; Table 1). Concentrations of T and 11KT were significantly elevated in women with Stage I, II, III and IV endometriosis compared to healthy controls, whereas 11KA4 was significantly reduced in women with all stages of endometriosis compared to healthy controls (Figure 1I-P). The greatest difference in androgen concentrations compared to healthy controls was in patients classified as Stage I. Median concentrations of T, 11KA4 and 11KT were modestly, inversely decreased with stage but overall, there was no distinct steroid signature associated with a given rASRM stage.

### Androgen metabolism is altered in endometriosis and biased towards production of 11-ketotestosterone

The correlation between serum androgen analytes was assessed in healthy controls and women with endometriosis using Pearson correlation (Figure 2A and B, Supplementary Table 3). In healthy controls, closely related metabolites exhibited a significant positive correlation. This included classic androgens; A4 and T (r=0.8, p=1.59E-12) and T and DHT (r=0.5, p=6.37E-06), as well as 11-oxygenated androgens; 11OHA4 and 11KA4 (r=0.6, p=8.2E-07) and 11OHT and 11KT (r=0.9, p=5.53E-20). In women with endometriosis, additional metabolites that were not correlated in healthy controls were found to be positively correlated. This included a significant positive correlation between classic and 11-oxygenated androgens, as well as a greater correlation amongst all 11-oxygenated androgen metabolites. This was characterised by DHEA, A4, T and DHT each being significantly positively correlated with all 11-oxygenated androgen metabolites (Figure 2A and B, Supplementary Table 3). Additionally, there was a significant positive correlation between 11KT and 11OHA4 (r=0.6, p=1.82E-19), 11KT and 11KA4 (r=0.7, p=7.68E-22), 11OHT and 11OHA4 (r=0.7, p=2.14E-24), and 11OHT and 11KA4 (r=0.6, p=3.94E-17) in women with endometriosis that was absent in healthy controls (Figure 2A and B, Supplementary Table 3).

**Figure 2.**
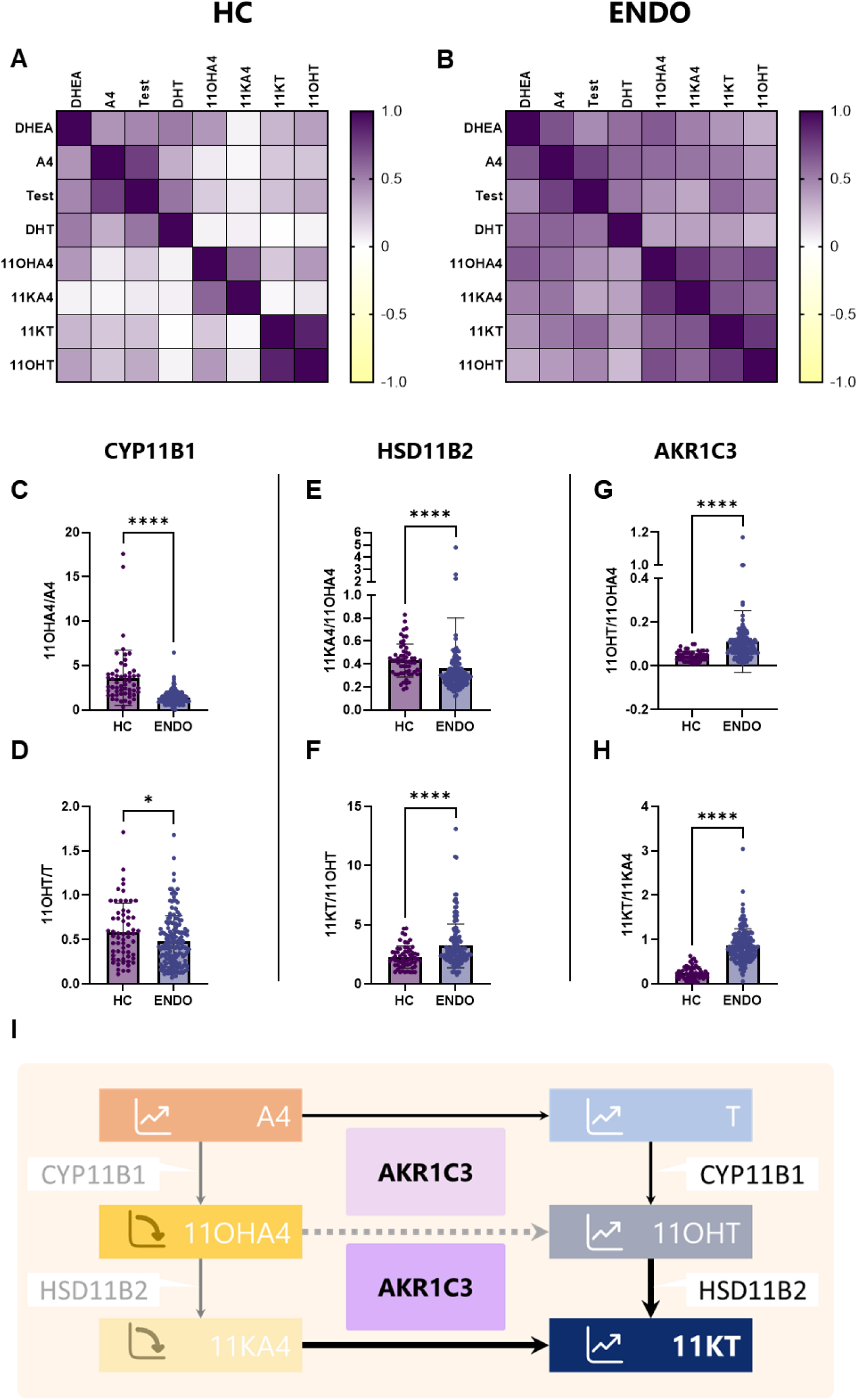
Endometriosis is associated with altered androgen metabolism characterised by increased production of 11KT. **A** Pearson correlation matrix of key analytes in HC demonstrates strong correlation between related steroids within relevant subgroupings including A4 and T, 11OHA4 and 11KA4 as well as 11KT and 11OHT. **B** In ENDO patients there was a strong correlation between both classic and 11-oxygenated androgen metabolites indicative of altered metabolism. Enzyme activity based on substrate to product ratios (product/substrate) were calculated based on measured serum concentrations in healthy control women (HC) and patients diagnosed with endometriosis (ENDO). The ratios required for sequential production of 11KT were assessed including CYP11B1 **(C, D)**, HSD11B2 **(E, F)** and AKR1C3 **(G, H)**. The ratios of 11OHA4/A4 **(C)**, 11OHT/T (**D)** and 11KA4/OHA4 **(E)** were significantly decreased in ENDO compared to HC. In contrast, the ratios of 11KT/11OHT **(F)**, 11OHT/11OHA4 **(G)**, 11KT/11KA4 **(H)** were significantly increased in ENDO compared to HC. Collectively these metabolism changes are associate with increased production of 11KT **(I)**. Statistical comparison Mann Whitney U test. *p<0.05, ****p<0.0001.

To investigate if these changes were indicative of altered metabolism in women with endometriosis, we estimated the activities of enzymes required for 11-oxygenated androgen production by calculating substrate to product ratios (Figure 2; 11-β-hydroxylase (CYP11B1), 11β-hydroxysteroid dehydrogenase type 2 (HSD11B2) and Aldo-keto reductase family 1 member C3 (AKR1C3)). CYP11B1 and HSD11B2 enzymes mediate interconversion and activation of 11-oxygenated androgens. The ratios of 11OHA4/A4 and 11OHT/T (mediated by CYP11B1) were significantly decreased in women with endometriosis (Figure 2C and D), consistent with decreased bioavailability of the precursors 11OHA4 and 11OHT. The ratio of 11KA4/11OHA4 (mediated by HSD11B2) was significantly decreased in women with endometriosis (Figure 2E, p<0.0001), consistent with decreased bioavailability of the precursor 11KA4. In contrast, the ratio of 11KT/11OHT was modestly increased between healthy controls and women with endometriosis (Figure 2F, p<0.0001) suggesting metabolism favouring production of 11KT. Ratios for the reciprocal inactivation of 11KA4 and 11KT, mediated by HSD11B1^28^, were also calculated. The ratio of 11OHA4/11KA4 was increased (Supplementary figure 1A, p<0.0002) and 11OHT/11KT controls was decreased (Supplementary figure 1B, p<0.0002) in women with endometriosis compared to healthy controls.

AKR1C3 is a multifunctional enzyme that plays a pivotal role in all pathways to androgen production including that of T, 11OHT and 11KT. The ratio of T/A4 (mediated by AKR1C3) did not differ between groups (Supplementary figure 1C), but other AKR1C3 activities determined by the ratios of 11OHT/11OHA4 and 11KT/11KA4 were significantly higher in women with endometriosis (Figure 2G and H; p<0.0001), suggestive of greater production of 11OHT and 11KT in women with endometriosis. Taken together, these data are consistent with a shift in metabolism towards production of 11KT (Figure 2I).

### Androgen concentrations can be used to discriminate between healthy controls and women with endometriosis

Receiver operating characteristic (ROC) curve analysis of serum androgen concentrations was performed to identify if specific androgen metabolites or ratios could be used to discriminate between healthy controls and women with endometriosis (Supplementary Table 4). The androgen analytes T, 11KA4 and 11KT exhibited best sensitivity and specificity with area under the receiver operating characteristic curve (AUC) values of 0.815 (p<0.001), 0.822 (p<0.001), and 0.815 (p<0.001) respectively (Figure 3A-C).

**Figure 3.**
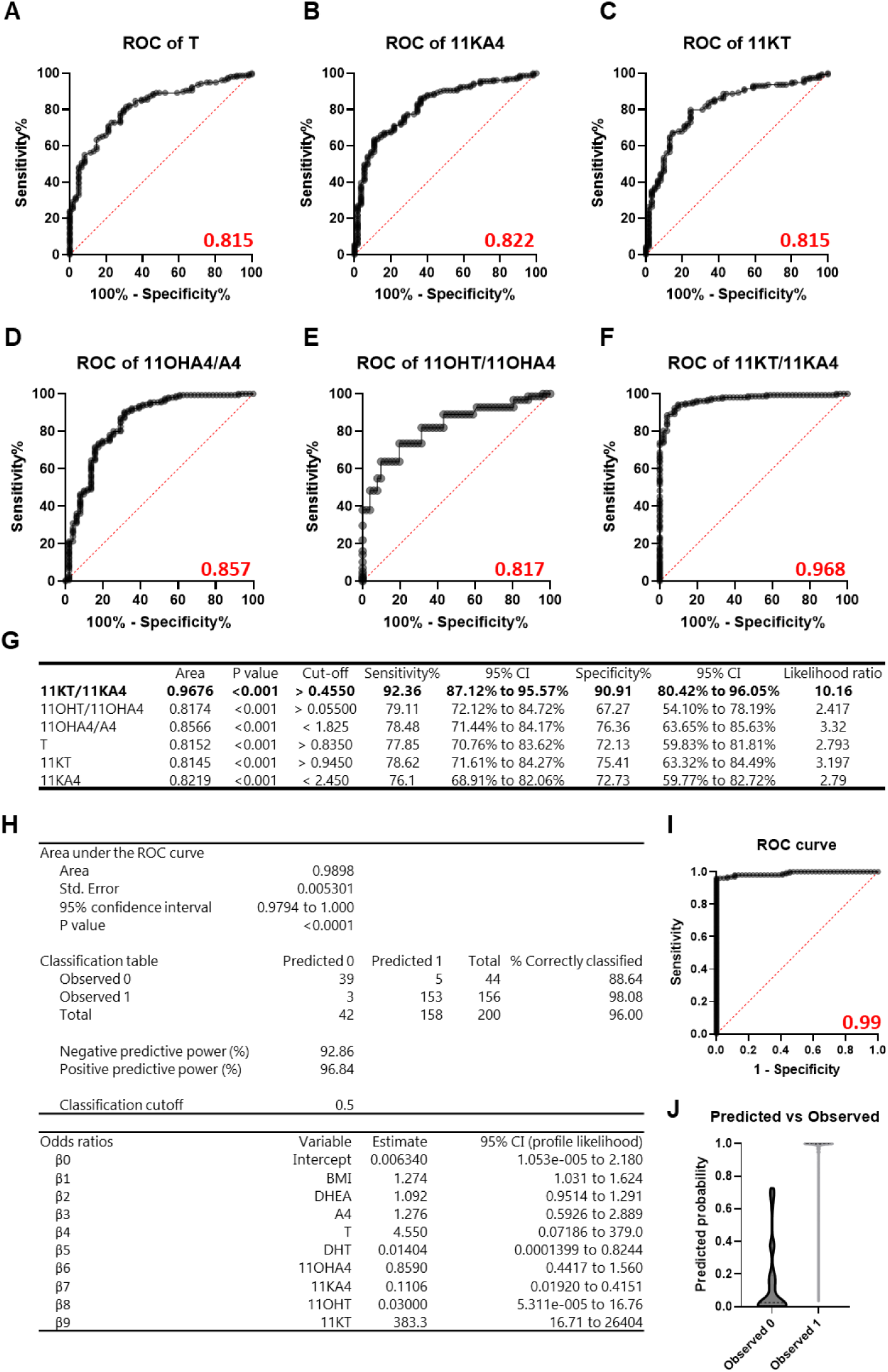
Assessing the utility of androgen concentrations for discrimination between HC and ENDO. ROC curve analysis of serum analyte concentrations **(A-C)** and ratios **(D-F)** was assessed (**G**). Robust discrimination between HC and ENDO was observed for single analytes; T (AUC 0.8152; p<0.001), 11KA4 (AUC 0.8145; p<0.001) and 11KT (AUC 0.8219; p<0.001), and for each ratio; 11OHA4/A4 (AUC 0.8556; p<0.001), 11OHT/11OHA4 (AUC 0.817; p<0.001), and 11KT/11KA4 (AUC 0.9676; p<0.001). Sensitibity, specificity and likelihood ratio were summarised and the ratio of 11KT/11KA4 provided the most robust discrimination between HC and ENDO **(G)**. **H** Multiple logistic regression modelling was used to fit a predictive model using androgen concentrations, Age and BMI as dependent variables and diagnosed endometriosis as the outcome variable. **I** ROC curve analysis of model demonstrated excellent discrimination between HC and ENDO with AUC of 0.99 (p<0.0001, CI 0.9794 to 1.000). **J** The predictive power of the model was calculated based on observed vs predicted outcomes. The negative predictive power of the model was 92.86% and the positive predictive power was 96.84%. Statistical comparison Mann Whitney U test. ****p<0.0001, ns – non significant.

ROC curve analysis of calculated enzyme ratios was also performed as these data have reduced inter-sample variability and provide a better indicator of the proposed altered metabolism found in women with endometriosis. Enzyme ratios associated with metabolism of 11-oxygenated androgens were assessed (Supplementary Table 4). The ratios 11OHA4/A4, 11OHT/11OHA4 and 11KT/11KA4 had best sensitivity and specificity with AUC values of 0.857 (p<0.001), 0.817 (p<0.001), and AUC of 0.968 (p<0.001) respectively (Figure 3D-F). The ratio of 11KT/11KA4 provided robust discrimination between healthy controls and women with endometriosis with a sensitivity of 92.36% and specificity of 90.91% and a likelihood ratio of 10.16 (Figure 3G). These data suggest that changes in androgen metabolism could be used as predictors of endometriosis diagnostic outcome.

### Development of a predictive model using supervised machine learning

To complement and extend these findings, androgen metabolome data were used to train a predictive model using multiple logistic regression. Androgen concentrations; DHEA, A4, T, DHT, 11OHA4, 11OHT and 11KT, were used as dependent variables and diagnosed endometriosis (as a binary) was the outcome variable. BMI was also included in the model to account for any independent effects of this variable (Figure 3H). ROC curve analysis demonstrated excellent discrimination between healthy controls and women with endometriosis with AUC of 0.99 (Figure 3I; p<0.0001, CI 0.9878 to 1.000) providing further improvement on discrimination informed by single analytes or ratios (Figure 3A-G). The predicted probability for outcome was calculated for each sample using a classification cut-off of 0.5. The negative predictive power of the multiple logistic regression model was 92.86% and positive predictive power 96.84% (Figure 3H and J) consistent with a robust and accurate model for predicting diagnostic outcome in endometriosis.

### Train and validation using a refined statistical model has robust positive predictive power for diagnostic outcome

To assess how a predictive model could perform when blinded to outcome, the whole data set was partitioned into train and validation groups. The training set contained dependent and outcome variables for an arbitrary subset of healthy controls (n=21) and women with endometriosis (n=21) and the remaining validation set contained dependent variables but was blinded to outcome variable (n=138). Comparative modelling was used initially to identify the most robust analytes to include in a refined predictive model (not shown). Multiple logistic regression was used to fit a predictive model using BMI, 11OHA4, 11KA4 and 11KT as dependent variables and diagnosed endometriosis as the outcome variable (Figure 4A, B). ROC curve analysis of the simplified model demonstrated excellent discrimination between healthy controls and women with endometriosis with AUC of 0.97 (Figure 4C; p<0.0001, CI 0.9124 to 1.000). The estimated negative predictive power of the model was 94.74% with a positive predictive power of 95.24% (Figure 4A and D).

**Figure 4.**
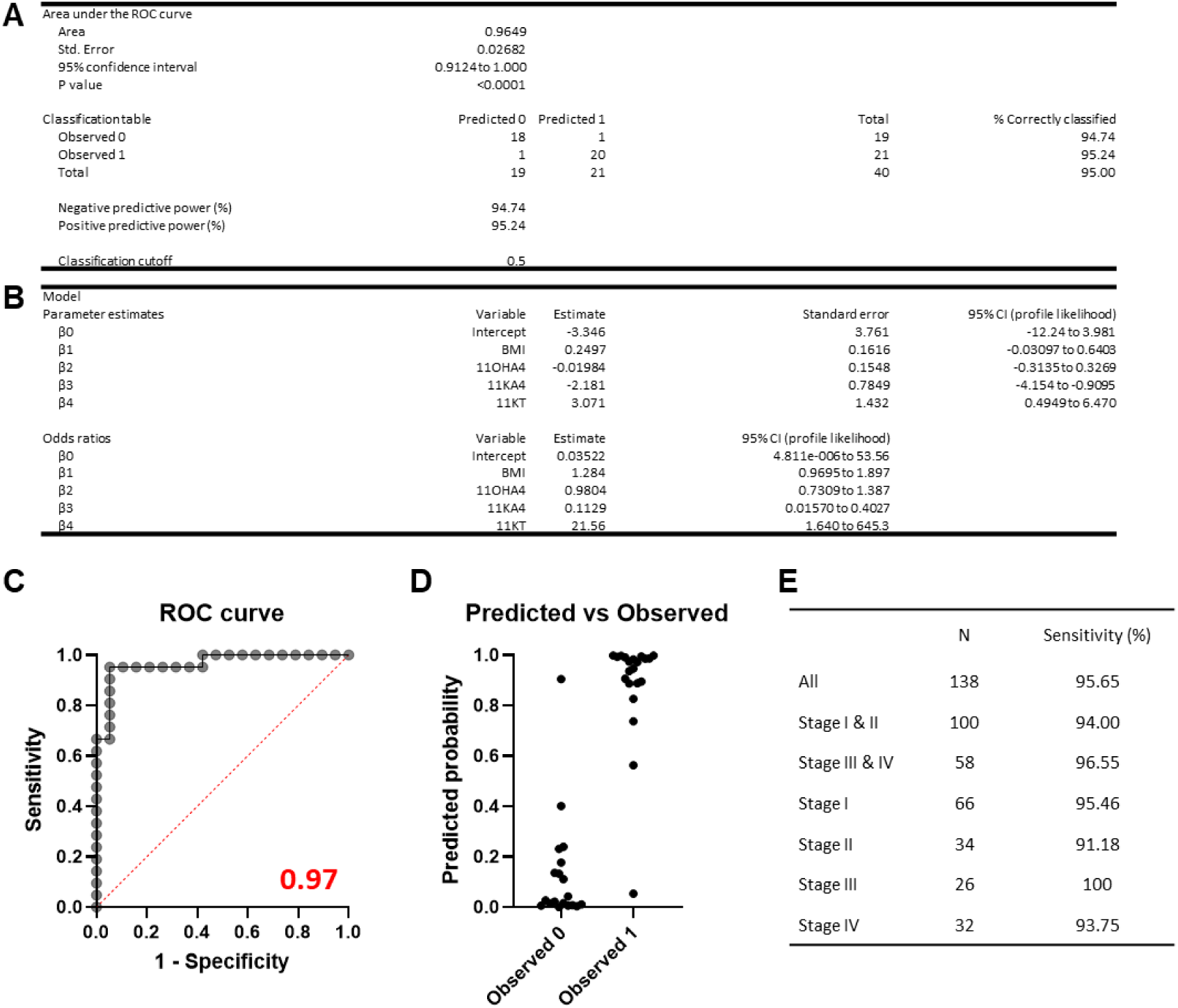
Actual performance of simplified predictive model for diagnostic status using androgen concentrations. The whole data set was partitioned into train and validation groups. The training set contained dependent and outcome variables for HC (n=21) and ENDO (n=21) and the validation set contained dependent variables but was blinded to outcome variable (n=138). Multiple logistic regression was used to fit a refined predictive model using BMI, 11OHA4, 11KA4 and 11KT as dependent variables and diagnosed endometriosis as the outcome variable (**A**, **B**). Samples with missing data were excluded as part of the regression analysis. **C** ROC curve analysis of the model demonstrated excellent discrimination between HC and ENDO with AUC of 0.965 (p<0.0001, CI 0.9124 to 1.000). The estimated negative predictive power of the model was 94.74% with a positive predictive power of 95.24%. (**A**, **D**). The actual performance of the model was assessed using the remaining data with outcome blinded (validation set). **E** The sensitivity for ENDO was 95.65% with accurate prediction observed in all disease stage stratifications; including grouped Minimal/Mild (Stage I + II), Moderate/Severe (Stage III + IV) or by individual rASRM stage (I-IV).

The actual performance of the model was assessed using the remaining data with outcome blinded (validation set) and included separate performance estimates according to stratification by rASRM stage (Figure 4E). The predicted outcome was calculated for each sample, using a classification cut-off of 0.5. In the validation dataset, the sensitivity of the refined model was 95.65% with accurate prediction observed for all disease stage stratifications (Figure 4E); including grouped minimal/mild ENDO (Stage I + II; 94% sensitivity), moderate/severe ENDO (Stage III + IV; 96.55% sensitivity) or by individual rASRM stage (I-IV; range 91.2-100% sensitivity). Consistent with univariate analysis, 11KT had a large odds ratio estimate, indicating the importance of this variable in determining diagnostic outcome. These data confirm the utility of using androgen concentrations to predict diagnostic outcome, with a refined model accurately identifying >95% of women with endometriosis in a blinded cohort.

## Discussion

In summary, we sought to define the androgen metabolome in endometriosis and investigate its potential utility as a diagnostic approach. We discovered a disease-specific hormone signature that identifies endometriosis with high specificity and sensitivity. This is characterised by increased concentrations of classic and 11-oxygenated androgens in serum, reframing endometriosis as an androgen-dependent disorder. Elevated androgens were associated with changes in activities of androgen synthesis enzymes, including AKR1C3, indicative of altered metabolism that results in increased production of 11KT in women with endometriosis. When used alone or in combination with multiple logistic regression, androgen measurements provided a strong predictive performance for endometriosis using both comprehensive and targeted analyte statistical models. When data were partitioned into train and validation cohorts, a refined model using only 11OHA4, 11KA4 and 11KT in combination with BMI was sufficient to correctly identify >95% of women with endometriosis in a blinded sample set. These data provide a significant breakthrough in the search for endometriosis blood biomarkers and place 11-oxygenated androgens at the cornerstone of a new diagnostic approach for endometriosis.

Several blood biomarkers for endometriosis have been proposed, such as the glycoprotein CA-125, inflammatory cytokines (e.g. IL-6, IL-8, TNF-α and MCP-1 [CCL2]), as well as the pro-angiogenic factor VEGF. Although these markers have been found to be elevated in women with endometriosis, they lack specificity and have not been validated in larger studies. A recent multi-centre study that assessed 54 blood-based biomarkers in samples from 919 women with endometriosis found CA-125 offered best discrimination between endometriosis cases and pathology-free symptomatic controls. However, the AUC for CA-125 in this comparison was 0.645 and thus had limited accuracy as a diagnostic biomarker ^29^. Critically, CA-125 is elevated in other conditions including ovarian and endometrial cancers, further limiting its utility as a diagnostic biomarker in endometriosis ^30,31^. The use of CA-125 as a diagnostic or screening tool is not recommended within the European Society of Human Reproduction and Embryology endometriosis guidelines ^32^.

Emerging results from a proteomic screen of serum samples from a large cohort of 805 patients showed promise using a combination of ten serum protein biomarkers with strong predictive accuracy for the diagnosis of endometriosis, particularly those with severe endometriosis (Stage IV) ^18^. Further studies are needed to establish if these candidate markers are specific to endometriosis and if their expression is affected by age, menstrual cycle stage or hormone treatment. Similarly, an undisclosed panel of 109 salivary miRNAs were reported to detect endometriosis with high sensitivity (96.2%) and specificity (95.1%) in an interim validation study, but whether these markers are variable across populations and endometriosis subtypes is not yet known ^33^.

An ideal biomarker for endometriosis would need to overcome the limitations of current approaches and should have high sensitivity and specificity, reflect endometriosis pathophysiology, be consistent across subtypes and not vary by menstrual cycle stage or with hormone treatment. Based on our results, 11-oxygenated androgens represent a significant improvement on other endometriosis biomarkers as they fulfil all of these key criteria. To the best of our knowledge, our study is the first to profile 11-oxygenated androgens in endometriosis. However, larger and more representative cohorts will be needed to understand if this profiling is generalisable across different demographics and endometriosis subtypes. The participants in our study were mostly of European ethnicity and future studies should ensure broader representation and inclusion to limit bias in this factor. Comparisons to other endocrine disorders will also be important to understand distinct features of endometriosis. Although there are major differences between the androgen analyte concentrations in endometriosis in our study and those previously published for PCOS, such as increased rather than decreased concentrations of 11OHA4 and 11KA4 in PCOS, an integrated analysis of both disorders would provide the greatest resolution on their distinct hormone profiles. We have profiled samples from all stages of endometriosis, but our study cohort was more representative of patients with minimal/mild disease (Stage I and II; 62.9% of cohort) rather than moderate/severe disease (Stage II and IV; 37.1% of cohort), and predominantly presented with pain rather than infertility symptoms. We did not detect a distinct hormone profile associated with endometriosis subtypes but it is possible that naturally occurring groups within the data could be identified using cluster analysis approaches in larger datasets. This could add further resolution to existing staging and subtyping methods used in endometriosis. This is important because current disease staging approaches do not correlate with severity of symptoms ^12^ which in part compounds the diagnostic challenges associated with endometriosis.

We found no correlation between androgen concentrations and pain scores in women with endometriosis but the majority of patients in our cohort presented with pelvic pain which may bias interpretation. Androgens play a complex role in pain perception in women with endometriosis that is yet to be fully defined. An inverse correlation between measures of chronic pain and androgen levels has been reported in women with dysmenorrhea but this study did not directly assess women with endometriosis ^34^. Notably, in our study we measured multiple androgen analytes in women with laparoscopically-confirmed endometriosis and correlated these with pain scores based on the pain domain of the EHP-30 questionnaire: a reliable and validated questionnaire for assessing the health-related quality of life in women with endometriosis ^35^. This direct comparison found no significant correlation between androgens and pain in women with endometriosis.

Although androgens did not correlate with endometriosis symptoms or subtypes, we believe that they may have a direct role in endometriosis pathogenesis. Androgens can regulate processes associated with formation of endometriosis lesions and the androgen receptor has been identified as an endometriosis-associated transcription factor ^36^. 11KT has similar potency to T in activating androgen-dependent processes ^25^ and was detected at higher concentrations than T in women with endometriosis in the current study. We propose that androgen excess in endometriosis drives pathophysiology through direct regulation of androgen-dependent processes (via T and 11KT) in endometrial and endometriotic tissues and by acting as obligate substrates for aromatase in lesions (via A4 and T) to fuel estrogenic processes. Changes to androgen metabolism may also have systemic effects that increase risk of lesion formation over the life course, including changes to innate and adaptive immune cell function and inflammation as reported in other contexts of androgen excess such as PCOS ^37^. This may explain how retrograde menstruation can lead to lesion formation in some women but not others.

Concentrations of 11KT are not diminished by oral contraceptives or gonadotropin-releasing hormone analogue-mediated ovarian suppression ^27^. If 11KT is driving endometriosis pathogenesis, this may explain why these widely used medical treatments for endometriosis-associated pain are ineffective in some women ^27^. Previously, the synthetic androgen danazol was found to be effective in the medical management of endometriosis-associated pain ^38^ but its use was stopped due to unacceptable masculinizing side-effects. The mechanism of action of danazol in endometriosis has not been fully characterised but it has been shown to inhibit adrenal androgen synthesis via suppression of androgen metabolising enzyme activity in other contexts ^39^. We have found that adrenal androgen production was a key feature of endometriosis; as characterised by altered activities of HSD11B2, HSD11B1 and AKR1C3 and the enhanced biosynthesis of 11KT. Thus, the efficacy of danazol in endometriosis may be due to its action on androgen metabolism, which reframes our perception of the pharmacodynamics of this drug. Although danazol was poorly tolerated, these results support revisiting danazol (e.g. administered by a local delivery system) or a more selective derivative as a therapeutic approach in endometriosis.

11KT is now considered the most biologically relevant androgen in women ^24^ and we have identified that endometriosis is associated with 11KT excess. These data highlight the importance of androgens in endometriosis, both as a fundamental feature of the pathophysiology and as a potential disease driver. When combined with predictive modelling androgen metabolite concentrations can be used to robustly distinguish between healthy controls and women with endometriosis. Collectively, these data signify a paradigm shift in our understanding of endometriosis and represent a unique opportunity to develop rapid, non-surgical approaches for endometriosis diagnosis and management.

## Data Availability

All data produced in the present study are available upon reasonable request to the authors

## Acknowledgements

This work was supported by the Wellcome Trust (Fellowship 220656/Z/20/Z to DAG, Investigator Award 209492/Z/17/Z to WA), Medical Research Council (Grant MRC/IAA/002 to DAG, program grant MR/N024524/1 to PTKS, and program grant MC_UP_1605/15 to WA) and the Institute for Regeneration and Repair Innovators award (funded by WT ITPA to DAG). Research conducted by the Edinburgh EXPPECT group that contributed to collection of biospecimens has been supported by grants from the MRC, Wellcome Trust, NIHR, CSO, and Wellbeing of Women. We thank members of the EXPPECT Edinburgh group, Frances Collins and Ann Doust, for support with participant recruitment and biosample collection. For the purpose of open access, the author has applied a Creative Commons Attribution (CC BY) licence to any Author Accepted Manuscript version arising from this submission.

## Materials and Methods

### Study approval

Written informed consent was obtained prior to study participation from healthy volunteers identified through advertisement. Ethical approval was granted by the Science, Technology, Engineering and Mathematics Ethical Review Committees of the University of Birmingham, UK (ERN_17-0494, ERN_17-0494B).

Written informed consent was obtained from all endometriosis study participants prior to surgery. Ethical approval was granted by the Lothian Research Ethics Committee (LREC 11/AL/0376), South Central-Hampshire A research ethics committee (IRAS:237815 REC reference 19/SC/0449) and Wales REC 6 A research ethics committee (IRAS 268806; REC ref: 19/WA/0271). Methods were carried out in accordance with Local Tissue Governance guidelines and international EPHect guidelines (https://endometriosisfoundation.org/ephect/).

### Patient cohorts

Inclusion criteria for healthy controls (HC) were pre-menopausal women aged 18 years or above. A standardized questionnaire was used to record demographic data including age and BMI; the use of hormonal contraceptives and menopausal status were recorded. Healthy control data were selected from a broader cohort of participants as detailed in Schiffer et al. to match age range of patient cohort in the current study ^27^.

Eligible participants were women with chronic pelvic pain (aged 18–50 years) of >3 months duration who were undergoing diagnostic laparoscopy for suspected endometriosis in NHS Lothian. Pelvic pain was defined as pain located within the true pelvis (between and below the anterior iliac crests). Participants at the Liverpool Women’s Hospital were those undergoing diagnostic laparoscopy for suspected endometriosis that were recruited from endometriosis or general gynaecology clinics. Diagnostic outcome, age, BMI, menstrual cycle stage and hormone status were obtained and recorded along with other key clinical data. Diagnosis of endometriosis was confirmed macroscopically at laparoscopy (ENDO). Endometriosis was subsequently classified according to the revised scoring system of the American Society for Reproductive Medicine (rASRM) which ranges from Stage I to Stage IV based on the location and extent of lesions, as well as adhesions, visualized at surgery ^11^.

Exclusion criteria were any acute or chronic disease affecting steroid biosynthesis or metabolism (including polycystic ovarian syndrome; PCOS) and the intake of any medication known to interfere with steroid biosynthesis or metabolism. Participants using hormonal contraceptives (combined oral contraceptives, contraceptive depot injection, or implant) were excluded.

### Serum steroid analysis

All venous blood samples were collected in the morning and, in the case of women with suspected endometriosis, prior to anaesthesia on the morning of surgery. All blood samples were collected in serum-separating tubes. After centrifugation, aspirated serum was aliquoted and stored at -80°C. Serum samples from women with endometriosis (n=159) and healthy controls (n=57) were profiled using a sensitive liquid chromatography tandem-mass spectrometry (LC-MS/MS) assay to simultaneously measure 21 steroid hormones/metabolites.

Serum steroids were quantified using a previously published and validated approach for multi-steroid profiling using liquid chromatography–tandem mass spectrometry (LC–MS/MS) assay ^40^. Briefly, 200 μL of serum was mixed with stable isotope-labelled internal standards and following protein precipitation with 50 μL acetonitrile samples were extracted by liquid–liquid extraction with 1 mL methyl tert-butyl ether (MTBE). The MTBE organic phase was removed, dried and reconstituted in 50/50 methanol/water. Steroids were chromatographically separated using a Phenomenex Luna Omega C18 column (1.6 µm, 100Å, 2.1 mm × 50 mm) and a water (0.1% formic acid)–methanol gradient. Ammonium fluoride was introduced by post-column infusion to aid ionisation. Steroids were quantified relative to a calibration series ranging from 0.02 to 250 ng/mL with inclusion of a blank calibration point. Analysis was performed on a Waters Xevo TQ-XS mass spectrometer using electrospray ionization in positive ion mode.

### Analyte key

Serum analytes were grouped as follows. Classic Androgens; dehydroepiandrosterone (DHEA), androstenedione (A4), testosterone (T) and dihydrostestosterone (DHT). 11-oxygenated androgens; 11β-hydroxyandrostenedione (11OHA4), 11β-hydroxytestosterone (11OHT), 11-ketoandrostenedione (11KA4) and 11-ketotestosterone (11KT).

### Statistical analysis

Statistical analysis was performed using GraphPad Prism 10. Undetected steroid concentrations or data below the LLOQ were replaced by 0.5×LLOQ for statistical purposes. Data were summarized using mean ± SD, presented for healthy controls (N=57) and women with endometriosis (N=159) or as median ± interquartile range in tables, as indicated. T test was used to compare the difference in the means of the two groups. Two-way ANOVA was used to determine the significance between treatments in grouped data. Non-parametric testing was utilised where sample sizes were insufficient to confirm normality of data distribution; Mann–Whitney test to assess variance between two groups or Kruskal–Wallis test was used to assess differences between multiple groups. Criterion for significance was P < 0.05. Correlation between analytes was assessed using Pearson matrix. Discrimination between cohorts using metabolite concentrations or ratios was assessed by ROC curve analysis. Predictive modelling for multiple analytes was performed using multiple logistic regression.

