## Supplementary data for "Steroid Metabolome Profiling Identifies a Unique Androgen Hormone Signature Associated with Endometriosis"

1 Figure, 4 Tables

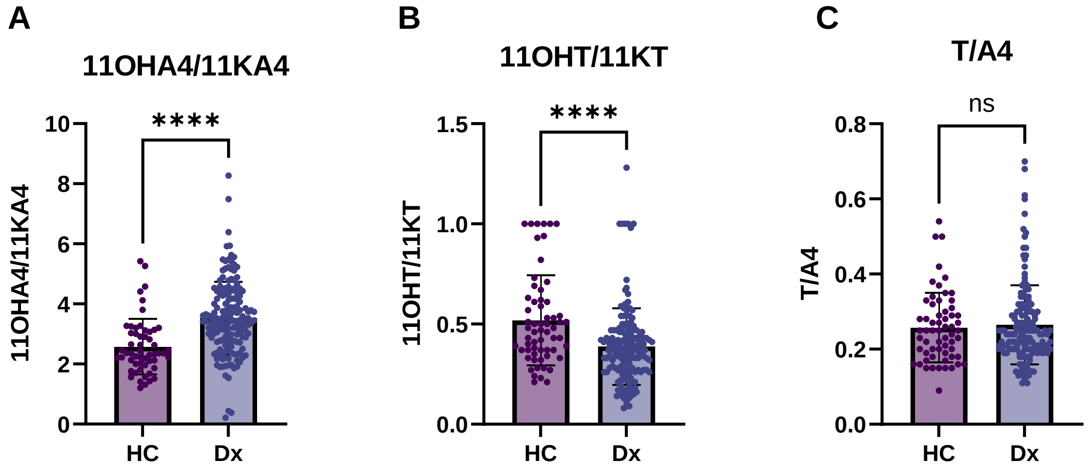


**Supplementary Figure 1.** Enzyme activity based on substrate to product ratios (product/substrate) were calculated based on measured serum concentrations in healthy control women (HC) and patients diagnosed with endometriosis (Dx). The ratios of 11OHA4/11KA4 (**A**, mediated by HSD11B1), 11OHT/11KT (**B**, mediated by HSD11B1), and T/A4 (**C**, mediated by both AKR1C3 and HSD11B1) were calculated. Statistical comparison Mann Whitney U test. ****p<0.0001, ns – non significant.

| | **HEALTHY CONTROLS** | | | **ENDOMETRIOSIS** | | |
|---|---|---|---|---|---|---|
| | Proliferative (*n*=19) | Secretory (*n*=22) | All (*n*=57) | Proliferative (*n*=75) | Secretory (*n*=67) | All (*n*=159) |
| AGE | 33.00 (23-42) | 30.50 (24-48) | 30 (22-48) | 33.00 (19-50) | 31.00 (19-45) | 32.00 (19-50) |
| BMI | 22.04 (17.3-37.3) | 22.77 (18.8-32.3) | 22.04 (17.3-37.3) | 26.00 (18.0-62.0) | 24.50 (18.0-40.0) | **25.00 (18.0-62.0) ***** |
| DHEA | 7.5 (5.30-9.950) | 11.38 (7.54-16.15) | 9.6 (6.02-14.63) | 21.10 (12.20-34.80) | 19.05 (12.60-32.38) | **20.80 (12.23-32.48) ****** |
| A4 | 2.110 (1.570-3.830) | 3.050 (1.985-4.050) | 2.830 (1.760-4.010) | 4.660 (3.370-6.330) | 4.860 (3.210-7.370) | **4.715 (3.370-6.585) ****** |
| TEST | 0.5900 (0.4900-0.7000) | 0.6250 (0.4900-0.8975) | 0.6300 (0.5000-0.9300) | 1.200 (0.968-1.435) | 1.080 (0.843-1.525) | **1.150 (0.860-1.490) ****** |
| DHT | 0.1700 (0.1700-0.5000) | 0.3750 (0.1700-0.4550) | 0.1700 (0.1700-0.5000) | 0.420 (0.170-0.580) | 0.450 (0.170-0.660) | 0.4000 (0.1700-0.5975) |
| 11OHA4 | 6.060 (4.525-8.538) | 6.750 (5.350-10.03) | 7.260 (5.350-10.12) | 5.570 (3.540-8.360) | 5.625 (3.800-7.743) | **5.670 (3.795-8.205) **** |
| 11KA4 | 3.250 (1.833-3.870) | 3.080 (2.280-3.750) | 3.180 (2.280-3.750) | 1.560 (1.080-2.570) | 1.725 (1.123-2.385) | **1.640 (1.113-2.385) ****** |
| 11KT | 0.6200 (0.3800-0.9200) | 0.7150 (0.4450-0.9200) | 0.6900 (0.4350-0.9650) | 1.330 (0.950-1.800) | 1.275 (1.000-1.845) | **1.300 (0.985-1.800) ****** |
| 11OHT | 0.1700 (0.1700-0.4200) | 0.3700 (0.1700-0.4975) | 0.3600 (0.1700-0.5200) | 0.540 (0.170-0.720) | 0.475 (0.343-0.680) | **0.5100 (0.170-0.715) ****** |
| AN | 0.3500 (0.3500-0.8200) | 0.3500 (0.3500-0.9775) | 0.3500 (0.3500-1.010) | 1.040 (0.350-1.820) | 1.380 (0.870-1.958) | **1.210 (0.350-1.870) ****** |
| 5ADIONE | <0.2600 (0.260-0.260) | <0.2600 (0.260-0.260) | <0.2600 (0.260-0.260) | 0.260 (0.260-0.260) | 0.260 (0.260-0.260) | **0.260 (0.260-0.260) **** |

**Supplementary Table 1.** Menstrual cycle stage- and endometriosis-specific serum steroid concentrations (median and 25th-75th centile range) of classic and 11-oxygenated androgens in healthy controls and endometriosis patients. Dehydroepiandrosterone (DHEA), androstenedione (A4), testosterone (T) and dihydrostestosterone (DHT). Alternative androgen pathway metabolites; androsterone (An) and androstanedione (5adione). 11-oxygenated androgens; 11β-hydroxyandrostenedione (11OHA4), 11β-hydroxytestosterone (11OHT), 11-ketoandrostenedione (11KA4) and 11-ketotestosterone (11KT).

| Age | DHEA | A4 | Test | DHT | 11OHA4 | 11KA4 | 11OHT | 11KT | An | Adione |
|---|---|---|---|---|---|---|---|---|---|---|
| **r** | -0.06969 | -0.05452 | 0.03376 | -0.08096 | -0.006826 | -0.07828 | 0.1267 | 0.02891 | -0.08280 | 0.03640 |
| **95% CI** | -0.2016 to 0.06471 | -0.1870 to 0.07987 | -0.1008 to 0.1671 | -0.2125 to 0.05342 | -0.1424 to 0.1290 | -0.2117 to 0.05805 | -0.007192 to 0.2562 | -0.1053 to 0.1621 | -0.2142 to 0.05158 | -0.09788 to 0.1694 |
| **R squared** | 0.004857 | 0.002972 | 0.001140 | 0.006554 | 4.660e-005 | 0.006127 | 0.01606 | 0.0008355 | 0.006856 | 0.001325 |
| **P value** | | | | | | | | | | |
| **P (two-tailed)** | 0.3091 | 0.4264 | 0.6233 | 0.2372 | 0.9219 | 0.2599 | 0.0636 | 0.6734 | 0.2266 | 0.5955 |

| BMI | DHEA | A4 | Test | DHT | 11OHA4 | 11KA4 | 11OHT | 11KT | An | Adione |
|---|---|---|---|---|---|---|---|---|---|---|
| **r** | 0.003608 | 0.03314 | 0.02154 | -0.07497 | -0.1156 | -0.2029 | -0.03780 | -0.04479 | -0.1001 | -0.03355 |
| **95% CI** | -0.1322 to 0.1393 | -0.1030 to 0.1681 | -0.1148 to 0.1571 | -0.2086 to 0.06137 | -0.2493 to 0.02251 | -0.3313 to -0.06702 | -0.1726 to 0.09842 | -0.1794 to 0.09149 | -0.2327 to 0.03610 | -0.1685 to 0.1026 |
| **R squared** | 1.302e-005 | 0.001098 | 0.0004641 | 0.005620 | 0.01335 | 0.04115 | 0.001429 | 0.002006 | 0.01002 | 0.001125 |
| **P value** | | | | | | | | | | |
| **P (two-tailed)** | 0.9587 | 0.6338 | 0.7574 | 0.2807 | 0.1006 | 0.0037 | 0.5869 | 0.5196 | 0.1493 | 0.6297 |

| Pain | DHEA | A4 | Test | DHT | 11OHA4 | 11KA4 | 11OHT | 11KT | An | Adione |
|---|---|---|---|---|---|---|---|---|---|---|
| **r** | 0.07617 | 0.08049 | -0.03835 | 0.01248 | -0.02923 | -0.04049 | -0.009899 | -0.08582 | 0.1448 | 0.2011 |
| **95% CI** | -0.1093 to 0.2565 | -0.1050 to 0.2605 | -0.2215 to 0.1474 | -0.1718 to 0.1959 | -0.2120 to 0.1555 | -0.2227 to 0.1445 | -0.1935 to 0.1743 | -0.2655 to 0.09967 | -0.04016 to 0.3202 | 0.01786 to 0.3713 |
| **R squared** | 0.005802 | 0.006478 | 0.001470 | 0.0001558 | 0.0008546 | 0.001639 | 9.800e-005 | 0.007365 | 0.02097 | 0.04045 |
| **P value** | | | | | | | | | | |
| **P (two-tailed)** | 0.4205 | 0.3946 | 0.6868 | 0.8951 | 0.7575 | 0.6689 | 0.9167 | 0.3639 | 0.1242 | 0.0319 |

**Supplementary Table 2** – Pearson correlation between androgen concentrations and Age, BMI or Pain score based on the pain domain of the EHP-30.

| HC | DHEA | A4 | Test | DHT | 11OHA4 | 11KA4 | 11KT | 11OHT |
|---|---|---|---|---|---|---|---|---|
| DHEA | | 0.000757 | 0.00011 | 1.83E-05 | 0.001949 | 0.716472 | 0.025716 | 0.00268 |
| A4 | 0.000757 | | 1.59E-12 | 0.012164 | 0.565642 | 0.813796 | 0.09599 | 0.07682 |
| Test | 0.00011 | 1.59E-12 | | 6.37E-06 | 0.139509 | 0.551185 | 0.058295 | 0.008405 |
| DHT | 1.83E-05 | 0.012164 | 6.37E-06 | | 0.72801 | 0.648992 | 0.982902 | 0.735716 |
| 11OHA4 | 0.001949 | 0.565642 | 0.139509 | 0.72801 | | 8.2E-07 | 0.11511 | 0.002295 |
| 11KA4 | 0.716472 | 0.813796 | 0.551185 | 0.648992 | 8.2E-07 | | 0.840832 | 0.475534 |
| 11KT | 0.025716 | 0.09599 | 0.058295 | 0.982902 | 0.11511 | 0.840832 | | 5.53E-20 |
| 11OHT | 0.00268 | 0.07682 | 0.008405 | 0.735716 | 0.002295 | 0.475534 | 5.53E-20 | |

| ENDO | DHEA | A4 | Test | DHT | 11OHA4 | 11KA4 | 11KT | 11OHT |
|---|---|---|---|---|---|---|---|---|
| DHEA | | 1.05E-22 | 2.1E-09 | 3.52E-15 | 1.81E-20 | 2.77E-11 | 4.5E-08 | 4.95E-05 |
| A4 | 1.05E-22 | | 1.72E-29 | 9.75E-18 | 9.07E-16 | 1.73E-13 | 5.97E-13 | 3.43E-07 |
| Test | 2.1E-09 | 1.72E-29 | | 8.84E-14 | 1.24E-08 | 1.03E-05 | 2.75E-16 | 3.39E-10 |
| DHT | 3.52E-15 | 9.75E-18 | 8.84E-14 | | 1.91E-06 | 1.95E-06 | 4.74E-07 | 0.000542 |
| 11OHA4 | 1.81E-20 | 9.07E-16 | 1.24E-08 | 1.91E-06 | | 2.13E-36 | 1.82E-19 | 2.14E-24 |
| 11KA4 | 2.77E-11 | 1.73E-13 | 1.03E-05 | 1.95E-06 | 2.13E-36 | | 7.68E-22 | 3.94E-17 |
| 11KT | 4.5E-08 | 5.97E-13 | 2.75E-16 | 4.74E-07 | 1.82E-19 | 7.68E-22 | | 3.8E-35 |
| 11OHT | 4.95E-05 | 3.43E-07 | 3.39E-10 | 0.000542 | 2.14E-24 | 3.94E-17 | 3.8E-35 | |

**Supplementary Table 3** – P values for Pearson correlation matrix.

Table 1. ROC curve analysis of serum androgen concentrations and androgen ratios in healthy controls and endometriosis patients.

| ANALYTE | ROC AREA | STD. ERROR | 95% CI | P VALUE |
|---|---|---|---|---|
| DHEA | 0.7885 | 0.03247 | 0.7249 to 0.8522 | **<0.001** |
| A4 | 0.7694 | 0.03445 | 0.7018 to 0.8369 | **<0.001** |
| TEST | **0.8152** | 0.03013 | 0.7562 to 0.8743 | **<0.001** |
| DHT | 0.5722 | 0.04112 | 0.4916 to 0.6528 | 0.1 |
| 11OHA4 | 0.6093 | 0.04065 | 0.5297 to 0.6890 | 0.01 |
| 11KA4 | **0.8219** | 0.03165 | 0.7599 to 0.8839 | **<0.001** |
| 11KT | **0.8145** | 0.03163 | 0.7525 to 0.8765 | **<0.001** |
| 11OHT | 0.67 | 0.03862 | 0.5943 to 0.7457 | **<0.001** |
| A4/DHEA | 0.5776 | 0.04523 | 0.4889 to 0.6662 | 0.08 |
| T/A4 | 0.5089 | 0.04568 | 0.4194 to 0.5984 | 0.84 |
| DHT/T | 0.7101 | 0.04074 | 0.6303 to 0.7900 | **<0.001** |
| 11OHA4/A4 | **0.849** | 0.03466 | 0.7810 to 0.9169 | **<0.001** |
| 11OHT/11OHA4 | **0.8224** | 0.03015 | 0.7633 to 0.8815 | **<0.001** |
| 11KT/11KA4 | **0.9695** | 0.01075 | 0.9484 to 0.9905 | **<0.001** |
| 11OHT/T | 0.6063 | 0.04412 | 0.5198 to 0.6928 | 0.02 |
| 11KA4/11OHA4 | 0.7023 | 0.03891 | 0.6260 to 0.7785 | **<0.001** |
| 11KT/11OHT | 0.7269 | 0.03939 | 0.6497 to 0.8041 | **<0.001** |

**Supplementary table 4** – AUC values and statistics from RCO curve analysis of single analytes and ratios.
